# An SEIR Model with Contact Tracing and Age-Structured Social Mixing for COVID-19 outbreak

**DOI:** 10.1101/2020.07.05.20146647

**Authors:** Ali Teimouri

## Abstract

In December 2019 a severe acute respiratory syndrome now known as SARS-CoV-2 began to surge in Wuhan, China. The virus soon spread throughout the world to become a pandemic. Since the outbreak various measures were put in place to contain and control the spread, these interventions were mostly based on compartmental models in epidemiology with the main goal of controlling and monitoring the rate of the basic and effective reproduction number. In this paper, we propose an SEIR model where we incorporate contact tracing and age-structured social mixing. We show the explicit relation between contact tracing and social mixing and other relevant parameters of the proposed model. We derive a formula for the effective reproduction number which is expressed in terms of reported cases, tracing quantities and social mixing. We use this formula to determine the expectation value of the effective reproduction number in London, UK.

## 1 Introduction

COVID-19 is a fast-spreading infectious disease which is caused by the novel coronavirus SARS-COV-2 [1, 2]. The rapid rate of the spread and lack of preparation soon translated to a pandemic causing major disruption globally [3, 4]. From the early days of the spread, many well-established models were proposed to contain and control the outbreak. These models were mainly the extensions of the simple SIR model [5]-[11] where the population is being divided into susceptible, infected and recovered compartments. Various parameters can be incorporated into the models to study and analyse the effect of intervention methods and also to predict the spread of the infectious disease.

In this paper, we propose an SEIR model integrated with contact tracing and age-structured social mixing. Contact tracing is an important intervention method when it comes to controlling an infectious outbreak [12]. As the name suggests it involves tracing the contacts of a reported infectious individual and remove them into isolation if needed, so further spread would be prevented. There are numerous methods of contact tracing and this intervention strategy proved to be effective in controlling the COVID-19 outbreak in countries such as China, South Korea, Taiwan and Japan [14]-[16]. In this study, we adopt contact tracing appropriate to the characteristics of the COVID-19. It is now established that infected individuals with the COVID-19 are distinguished into two groups of asymptomatic and symptomatic [17]. The asymptomatic individuals are infectious individuals who show no symptoms while symptomatic individuals are those who show symptoms, and hence are reported and might need hospitalisation. To this end, our model traces the contact of those individuals who are symptomatic and reported. The rationale of the model is that if the the traced contacts show symptoms they will be removed into isolation, and if they do not show symptoms upon tracing they will be monitored by healthcare workers or via a mobile application for the recommended time of two weeks by the WHO [12].

Another feature that we integrate into our model is age-structured social mixing matrix. This component of the model is important when it comes to effectively classifying the risk within different age groups in a society. One of the known characteristics of the COVID-19 is that it affects mid-age and elderly people more severely than those who are in the young age group [18]. As a result, to implement interventions such as social-distancing within households or at workplaces, it is important to know the mixing between different age groups. In this work, we show the theoretical scaling effect of the social mixing matrix on the value of the effective reproduction number.

We begin by giving an overview of the SEIR model in section 2.1, we then incorporate contact tracing and social mixing respectively into our model in sections 2.2 and 2.3. We then analyse our results in section 3 and provide a deterministic formula for keeping the effective reproduction number below one. We calculate the change in the effective reproduction number when contact tracing and social mixing are involved. In section 3.1 we provide an example where we determine the value of the expected effective reproduction number for London, United Kingdom (UK).

## 2 Model and Methods

### 2.1 Toy SEIR model

We begin by considering a simple and adequate SEIR compartmental model which captures the essence of the spread of the COVID-19 through a homogenous population. In this model, the population *N* is the the sum of Susceptible (*S*), Exposed (*E*), Symptomatic (*I*_*s*_), Asymptomatic (*I*_*a*_) and Recovered (*R*) individuals (i.e. *N* = *S* + *E* + *I*_*s*_ + *I*_*a*_ + *R*). By definition, exposed individuals are those incubating the disease and will be infected by the symptomatic and asymptomatic individuals, symptomatic individuals are those who are infected, show symptoms and will be reported or hospitalised, asymptomatic infected individuals are those who have mild or no symptoms and hence will not be reported and do not require hospitalisation. Both infected groups will eventually move into the recovered compartment, which includes recovered and dead cases. The system of the ordinary differential equations (ODE) which describe the dynamics of this model is,

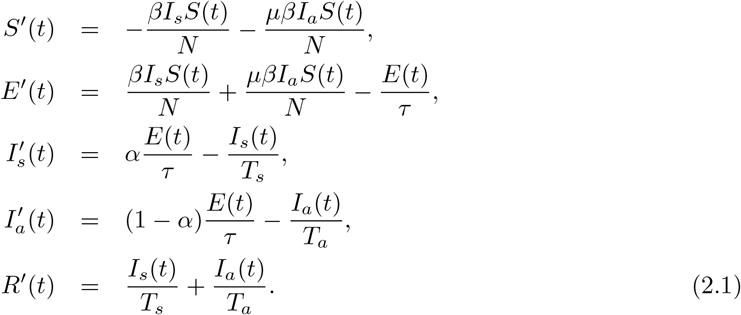

The ODEs are parameterised by *β* which is the transmission rate for infectious individuals, *μ* that is the multiplicative factor reducing the transmission rate of the asymptomatic infected patients, *τ*, the mean incubation period in days, *α*, the fraction of the reported/hospitalised cases, *T*_*s*_, the time from symptom onset until hospitalisation/isolation or reporting in days and *T*_*a*_, the mean infectious period in days for the asymptomatic individuals. Note that *T*_*s*_ is the same for the cases which are hospitalised, go to isolation or just being reported. The distinction of infected individuals into two groups is essential in this work as in reality only the symptomatic cased are being recorded and hence can trigger contact tracing [31]. Note that there are studies that take *T*_*s*_ and *T*_*a*_ as equal [32].

The basic reproduction number ℛ_0_ is defined as average number of secondary infections produced by an infected individual in a population where everyone is susceptible, this number is used to measure the transmission potential of a disease. In a realistic world a population can rarely be wholly susceptible. Some individuals can be immune, for instance due to prior infection. As a result, not all contacts will become infected and the average number of secondary cases per an infectious case will be lower than the basic reproduction number. To this end, the effective reproduction number ℛ_*e*_ is defined as the average number of secondary cases in a population made up of both susceptible and non-susceptible individuals. The effective reproduction number is proportional to the product of the basic reproduction number and the fraction of the host population that is susceptible. Using the next generation matrix method [19] the effective reproduction number, ℛ_*e*_, is calculated by

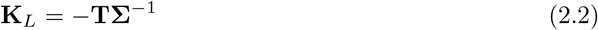

where the matrix **T** corresponds to transmissions and the matrix **Σ** corresponds to transitions. Note that all the epidemiological events that lead to new infections are incorporated in the model via **T** and the rest of the events via **Σ**, hence progress to either dead or recovery makes **Σ** invertible. We obtain **K**_*L*_ as,

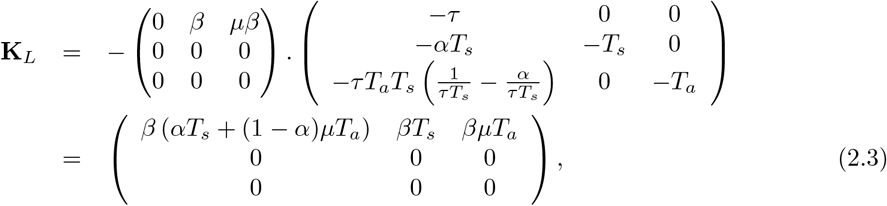

the dominant eigenvalue of this matrix is equal to the effective reproduction number,

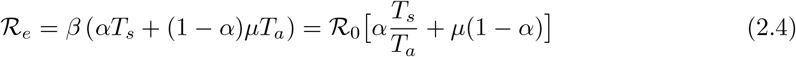

where ℛ_0_ = *βT*_*a*_ is defined as the basic reproduction number without cases being reported or hospitalised. We assumed that the population size *N* is constant over the short time of the outbreak and hence we absorb it into parameter *β* (i.e. we do not consider birth and death rates). Furthermore, we take the total population *N* = 1 so the results would be per-capita. We note that for *T*_*s*_ = *T*_*a*_ and *μ* = 1 the relation in the above equation would reduce to ℛ_*e*_ = ℛ_0_ where the assumption is that the basic reproduction number is only proportional to transmission rate and the mean infectious period. Namely this ratio would be the expected number of new infections from a single infection in a population where all individuals are susceptible.

### 2.2 SEIR model with contact tracing

To incorporate contact tracing into our model we follow the steps of [5]. Contact tracing in this manner is based on three elements, contact identification, contact listing and follow-up. This process begins once an infected (dead or alive) case is being reported, at this stage one lists the contacts of the reported person, then a team of contact tracers track the listed contacts (this could also be done via a mobile application). If the traced contacts show symptoms at the time of tracking they will be removed by isolation or quarantine. Otherwise, they will be instructed to update their status if they develop any symptoms over 14 days. The follow-up team would then look after the traced contacts during this period and will remove them if symptoms appear.

There are two assumptions to proceed with this model:

- Only the reported or hospitalised cases can trigger contact tracing.
- Contact of a reported patient are traced with probability *ϕ*. There is also a fixed delay *δ* between index case reporting and contact tracking. If the traced individuals are symptomatic at the time of contact, they will be removed immediately, otherwise they follow up the monitoring protocol.

Untraced contacts have probability (1 − *ϕ*), these are the cases that are missed and did not trigger tracing from the reported cases. We shall now define the cumulative number, at time *t*, of traced infected individuals who are in exposed or infectious group when first tracked,

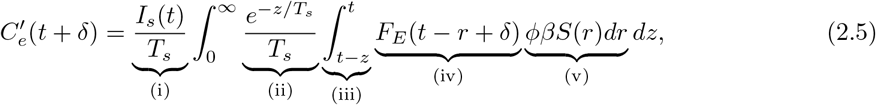

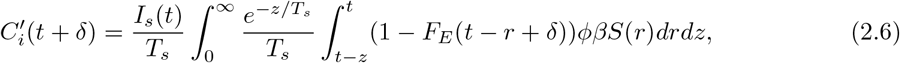

where:

i. is the rate of reporting of infectious individuals,
ii. is the reported case infectious period probability density,
iii. is the integral over infectious period of length *z*,
iv. is the probability still incubating *t* − *r* + *δ* units after infection and
v. is the traced transmissions infected at *r* ∈ [*t* − *z, t*].

We assumed that the incubation period cumulative distribution *F*_*E*_ is exponentially distributed. Since the average infectious period is short the the change in population size is negligible and one assumes that *S* is approximately constant over this period. Hence, the total number of the traced transmissions at time *t* + *δ* is,

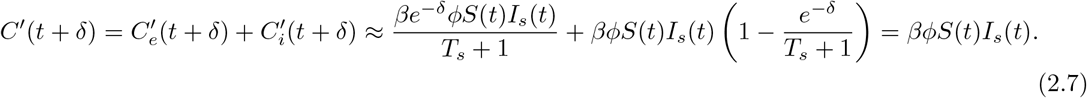

We now define four additional compartments, 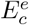 are the exposed individuals who will be traced and *incubating* when they were tracked, 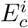 and *infectious* when tracked. 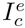 are the monitored (and under monitoring) infectious individuals who *have shown no symptoms* (while incubating) at the time of first contact, and 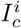 are the infectious individuals who should be removed as they were *found symptomatic* at the time of first contact.

The probability *p*_*e*_ of a traced contact being incubating when first tracked should depend on the mean infectious period *T*_*s*_, the delay between index case reporting and tracing *δ*, and the incubation period of the traced case *τ*, as a result:

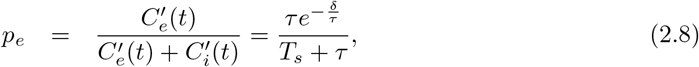

for *t* → *τ*. Note that we assumed the incubation period is exponentially distributed i.e. *F*_*E*_(*x*) = *e*^−*x/τ*^, and that *S*(*t*) is constant over the infectious period.

The average infectious period, prior to being tracked, for the fraction (1 − *p*_*e*_) individuals is defined by *T*_*i*_ and can be approximated by the expected value of the time after symptom onset when the contact is being tracked, 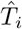,

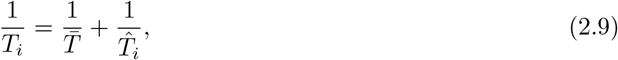

where 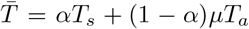 is the weighted average infectious period independent of contact tracing and

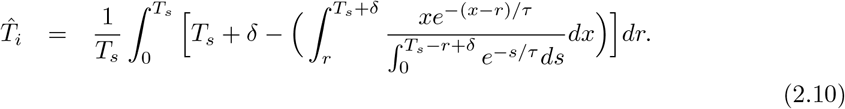

We assume monitored infectious individuals, 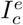 have transmissibility *β*_*m*_ and mean infection period *T*_*m*_ in days. The value of *β*_*m*_ and *T*_*m*_ directly depends on the efficiency of the contact tracing protocol. Note that for the infectious individuals, 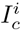, traced during their infectious period, we assign transmissibility *β* prior to tracking and mean infectious period *T*_*i*_.

Contact tracing which is triggered by individuals in 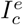 and 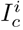 compartments counts as secondary tracing, tertiary tracing and so on and thus we call them *higher order tracing*. In contrast, the first order tracing are the contacts of the fraction *ϕ* who will be traced. We shall then define probabilities 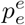 and 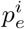 as probability of traced contact, infected by 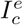 individuals, and probability of traced contact, infected by 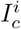 individuals, as

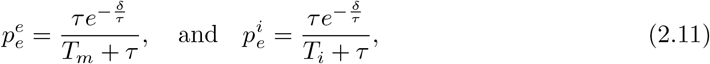

where we remind that *T*_*m*_ is the mean infectious period for the monitored infectious individuals, 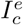, (time between symptom onset and isolation) and *T*_*i*_ is the mean infectious period (prior to being tracked) for the contact traced infectious individuals, 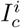. The modified SEIR model with contact tracing is,

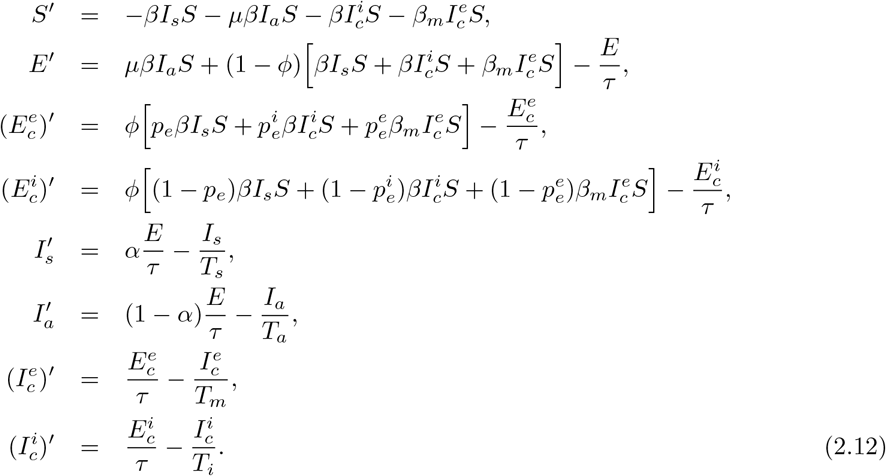

In the same manner as in the previous section the effective reproduction number can be obtained by,

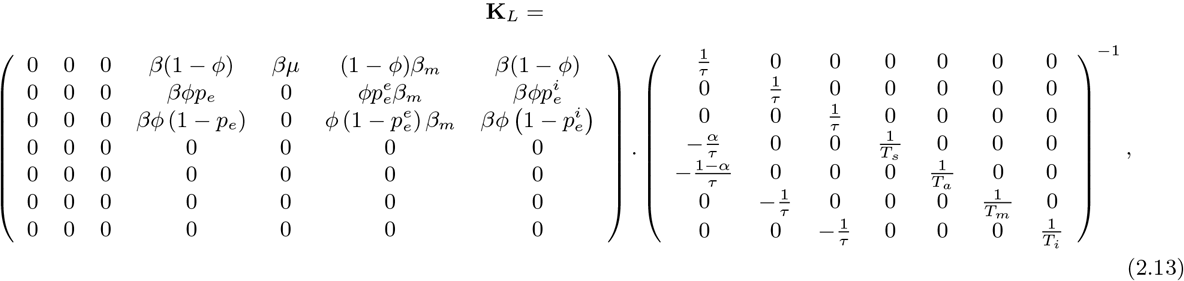

where the dominant eigenvalue of this matrix is,

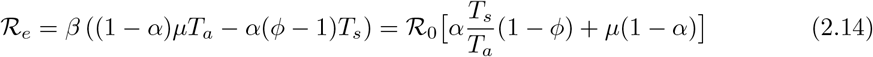

as before ℛ_0_ = *βT*_*a*_ is the basic reproduction number independent of *T*_*s*_. *α* is the fraction of untraced cases which are reported and *ϕ* is the fraction of reported cases which are traced. The main difference of the above equation and Eq.(2.4) is the probability *ϕ* which measures the likelihood of contacts of a reported case being traced. In the limit *ϕ* → 0 we recover Eq.(2.4) where there is no contact tracing.

### 2.3 SEIR model with contact tracing and social mixing

To take the social mixing into account we rewrite the transmission coefficient as the product of a transmission *β* times a constant matrix *C* whose element *C*_*λσ*_ measures the average number of physical contacts, on daily basis, among an individual in group age *λ* with an individual in group age *σ*. It should be noted that the probability of a susceptible individual, in age class *λ*, to come to contact with an infected individual in class *σ* is equivalent to the product of the contact rate *C*_*λσ*_ times the probability *I*_*σ*_ that individual in class *σ* is infected. Hence we can denote age groups appropriately via *λ* and *σ* and our model modifies to,

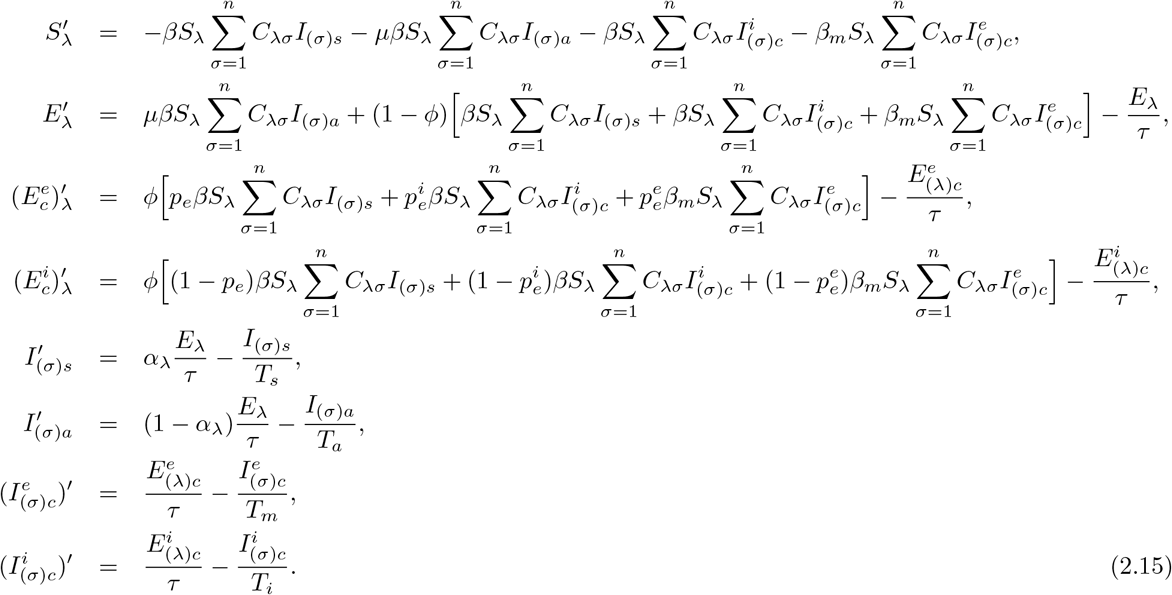

We note that *C*_*λσ*_ denotes the interaction between different age groups and hence it does not matter what groups of infected individuals it is coupled to. It should also be noted that the age structured mixing of individuals in the age group *λ* change their likelihood of being exposed to the virus given certain number of infectious people in the population. Furthermore, in our model we made a dependency between *α* and *λ* (i.e. *α* → *α*_*λ*_) due to the assumption that younger individuals are more likely to be asymptomatic and less infectious [9]. The effective reproduction number is now calculated as,

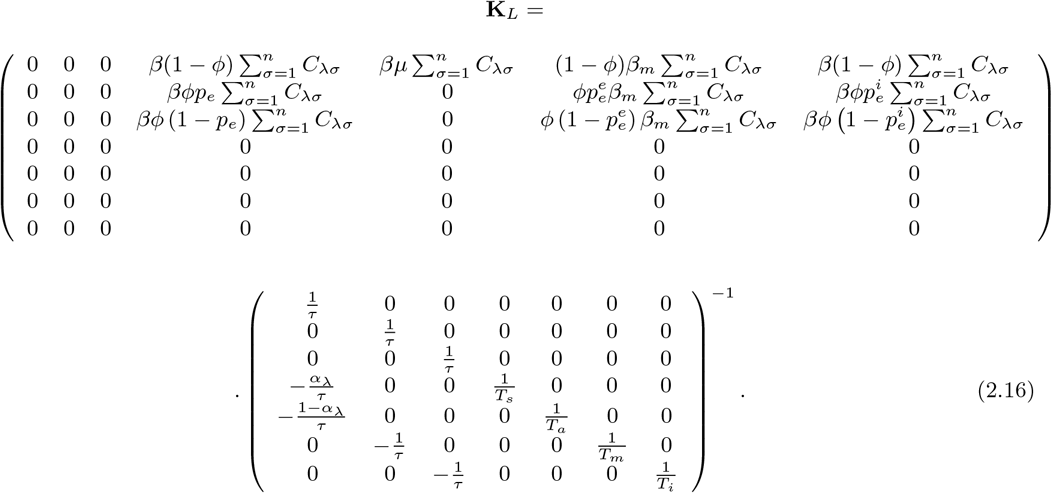

The dominant eigenvalue of this matrix is of the form,

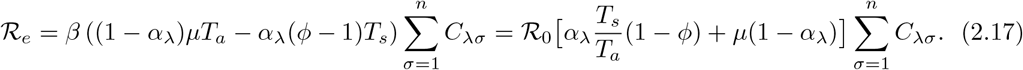

where *n ⩾* 1 is the number of age groups one wishes to consider. It can be seen from the formula above that the social mixing matrix *C*_*λσ*_ has a scaling effect on increasing or decreasing the effective reproduction number.

## 3. Results

*The impact of contact tracing on the effective reproduction number:* In order to keep the spread of the virus under control we need to keep the effective reproduction number below one. If we assume the same mean infectious period for both symptomatic and asymptomatic case (i.e. *T*_*s*_ ≈ *T*_*a*_) and also if we take the multiplicative factor *μ* = 1, we get,

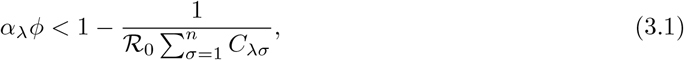

where *α*_*λ*_*ϕ* is the *critical* proportion of the total cases which need to be traced in different age groups in order for ℛ_*e*_ *<* 1.

Furthermore, the impact of contact tracing can be quantified by considering the relative change in the effective reproduction number. From the simple SEIR model let us recall the effective reproduction number and call it, 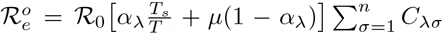, where we added the social mixing term appropriately. Then the relative change in ℛ_*e*_ with contact tracing would be of the form,

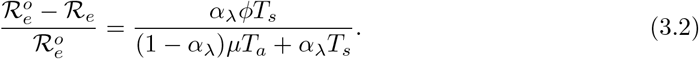

For *T*_*s*_ ≈ *T*_*a*_ and *μ* = 1 the relative change takes the critical form of *α*_*λ*_*ϕ* which is independent of the social mixing matrix *C*_*λσ*_ and is only proportional to the probability that contacts of a reported case being traced, *ϕ*, and fraction of untraced cases that are reported in a particular age group, *α*_*λ*_.

In literature [3], *μ* has a unconstrained distribution of *μ* ∼ uniform(0.2, 0.55). *α*_*λ*_ has a unconstrained distribution of *α*_*λ*_ ∼ uniform(0.1, 0.99). In Fig. 1 we plot the relative change for upper and lower values of *μ* while keeping *T*_*s*_ ≈ *T*_*a*_. Higher value of *μ* means higher transmission rate for asymptomatic individuals, as a result the relative change in the effective reproduction number with tracing is proportional to ∼ *α*_*λ*_*ϕ* for upper bound of *μ* while proportional to ∼ *ϕ* for lower bound of *μ*.

**Figure 1:**
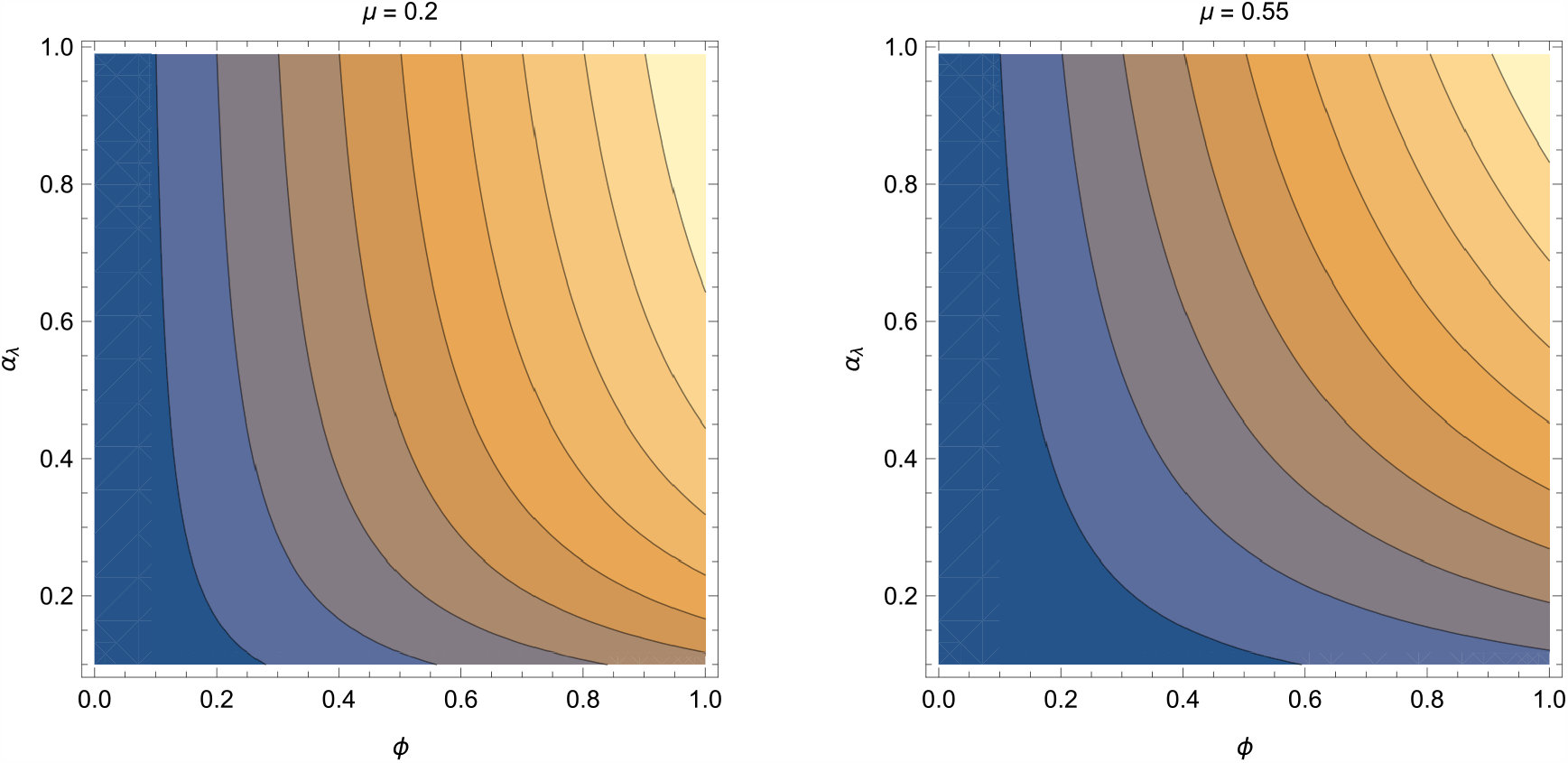
Relative change in the effective reproduction number, ℛ_*e*_, given by 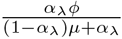. The plot varies the values of *α*_*λ*_ according to its distribution (i.e. *α*_*λ*_ ∼ uniform(0., 0.99)) and *ϕ* (where 0 ⩽*ϕ* ⩽ 1) for two values of *μ* = 0.2 and *μ* = 0.55.

*Imperfect tracing:* So far we assumed perfect tracing, that is to consider that traced individuals do not cause secondary infections due to an effective monitoring and tracking policies. In other words, we assumed transmission rate for monitored individuals is zero (i.e. *β*_*m*_ = 0). Let us now relax this condition and assume that *β*_*m*_ *>* 0, then the full dominant eigenvalue of (2.16) is,

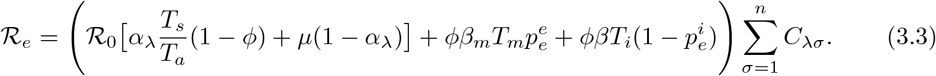

To capture the effect of the tracing let us assume 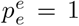 that is to consider that higher order traced contact is 100% infected by 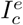 individual, let us also assume that 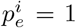 meaning that higher order traced contact is 100% infected by 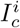 individual. In that case, the effective reproduction number reduces to,

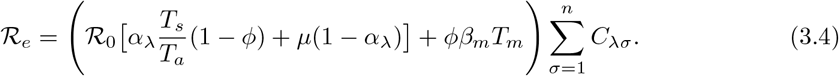

We remind that 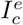 are the monitored infected individuals who have shown no symptoms upon contacting them and 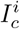 are the symptomatic infectious individuals who were traced and removed into isolation. It is now trivial that the efficiency of contact tracing has a linear contribution in the value of the effective reproduction number given by *ϕβ*_*m*_*T*_*m*_. The formula above shows that in order to have an effective monitoring policy in place the mean infectious period for monitored infectious individuals should be minimised. In other words, the time between symptom onset and isolation, *T*_*m*_ should be at its minimum.

*The impact of age-structured social mixing on the effective reproduction number:* To impose an effective distancing policy one needs to study the social mixing matrices that illustrate the average number of contacts between different age groups. Measuring the social mixing varies for different locations and could depend on the structure of households, schools and workplaces [9, 20]. In this paper, we are using the social mixing matrices constructed by [20]. The study by [20] conducted in the UK by surveying a sample of adults and recording their contact patterns after the British government put social distancing measures in place. In Fig. 2 we reconstructed the social mixing matrix from CoMIX study [20], there are six age groups participated in their study, namely 18-29, 30-39, 40-49, 50-59, 60-69, and 70+. As a result 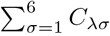. The CoMIX matrices show the average number of contacts between different age groups in different setups from the 24th of March 2020 one day after the lockdown was imposed in the UK. To control the spread of the virus one should monitor the number of contacts between each age group and minimise them when necessary. In the next section we use the mixing matrices in Fig. 2 to illustrate the effect of social mixing on the expected value of ℛ_*e*_.

**Figure 2:**
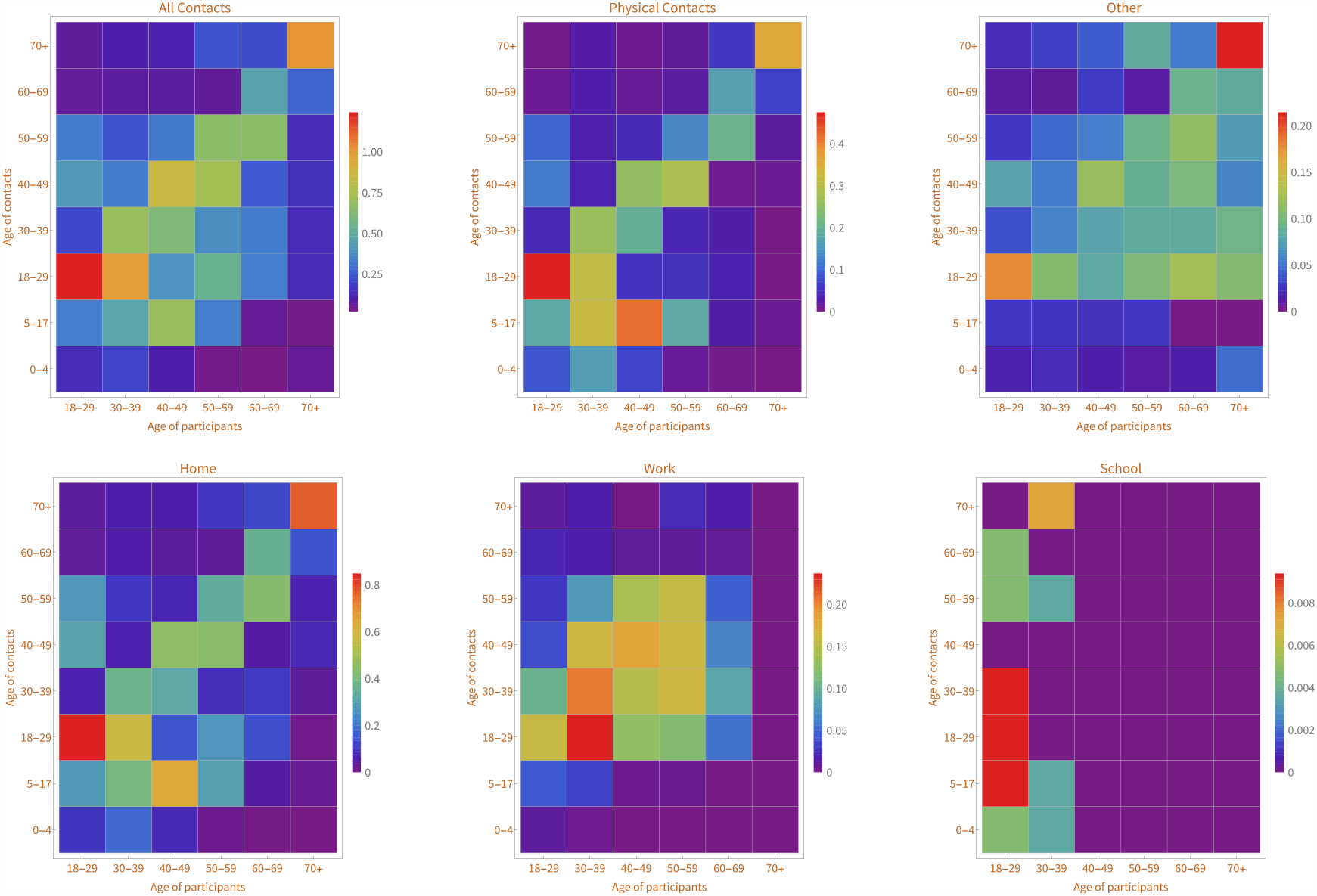
Reconstruction of CoMIX matrices as given in [20], each matrix represents a social setup and show the average number of contacts between different age groups after the lockdown was imposed in the UK on the 23rd of March 2020.

### 3.1 The expected value of the effective reproduction number in London

Let us recall Eq. (3.4) and rewrite it as:

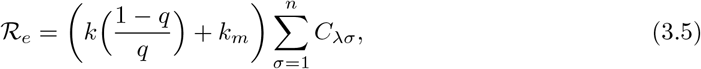

where we define *k* = *ϕ*ℛ = *ϕβT*_*s*_, in this definition ℛ is the basic reproduction number in the absence of contact tracing, note that this is different than ℛ_0_ which was defined earlier.

Moreover, let

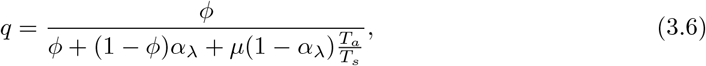

be the fraction of reported cases which are traced contacts. In fact, *q* can be interpreted as the conditional probability that a transmission is traced given a cases is being reported. Finally, *k*_*m*_ = *ϕ* ℛ_*m*_ = *ϕβ*_*m*_*T*_*m*_ where ℛ_*m*_ is the reproduction number of the contact traced / monitored individuals. Since *k* and *k*_*m*_ are the infected traced contacts identified per untraced and traced reported cases respectively, it is sensible to define 𝒞_*j*_ and 𝒯_*j*_ as the cumulative reported cases and traced-reported cases in week *j* respectively. We now define the average number of traced infected individuals identified in week *j* + *i* per untraced and traced reported case from the preceding week *j* as,

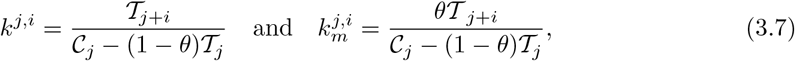

where 0 ⩽ *θ* ⩽ 1 is tracing/monitoring effectiveness, in other words, 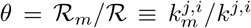. Furthermore, the fraction of reported cases that are traced in the week *j* + *i* is

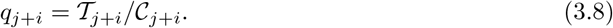

We can now write the expectation value of the effective reproduction number, given in week *j*, as

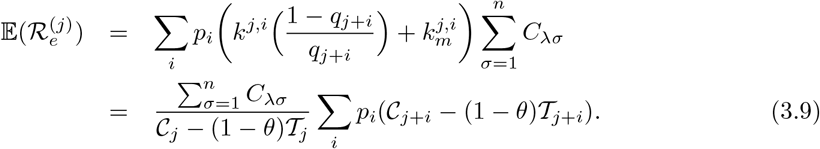

Above estimates 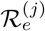 as the ratio of the sum of untraced and fraction of traced reported cases (*θ*), weighted by probabilities *p*_*i*_. The probabilities *p*_*i*_ are calculated using the serial interval of the COVID-19 that is estimated to be a normal distribution with mean of *μ* = 3.96 and standard deviation of *σ* = 4.75, [26], therefore on weekly basis we have: *p*_1_ = 𝕡(𝒩 *<* 7), *p*_2_ = 𝕡(7 *<* 𝒩 *<* 14) and so on. In perfect tracing (i.e. *θ* = 0) only weekly untraced reported cases (𝒞_*i*_ − 𝒯_*i*_) play role in the calculation of ℛ_*e*_ while for imperfect tracing (i.e. *θ* = 1) all the traced and untraced reported cases are weighted equally. As a result, one does not require specific data about the traced reported cases per week. Utilising *θ* in this manner could serve as a useful tool for sensitivity analysis and in the countries where obtaining the contact tracing data is difficult.

We are now equipped to calculate the expected value of the effective reproduction number for London. We begin by using the social mixing matrices given in Fig. 2, where there are six age groups in participants (*n* = 6) reporting their contact pattern with eight different age groups. We note that each matrix element is the average number of contacts between different age groups. As a result, the normalised sum of each matrix’s row gives the value of *C* for the particular age class and particular setup. Using the cumulative reported cases from [27] for London and Eq. (3.9) we estimated the expected value of ℛ_*e*_ - for nine weeks beginning from the 22nd of February 2020 - for different age groups in different setups in Fig. 3. Our results show that before the lockdown the mean of the effective reproduction number is 2.55 and after the lockdown is imposed the mean of the effective reproduction reduces to 0.77 over the weeks four to nine for all age groups and setups, this is comparable to results of [20] and [25] (when averaged on weekly basis).

**Figure 3:**
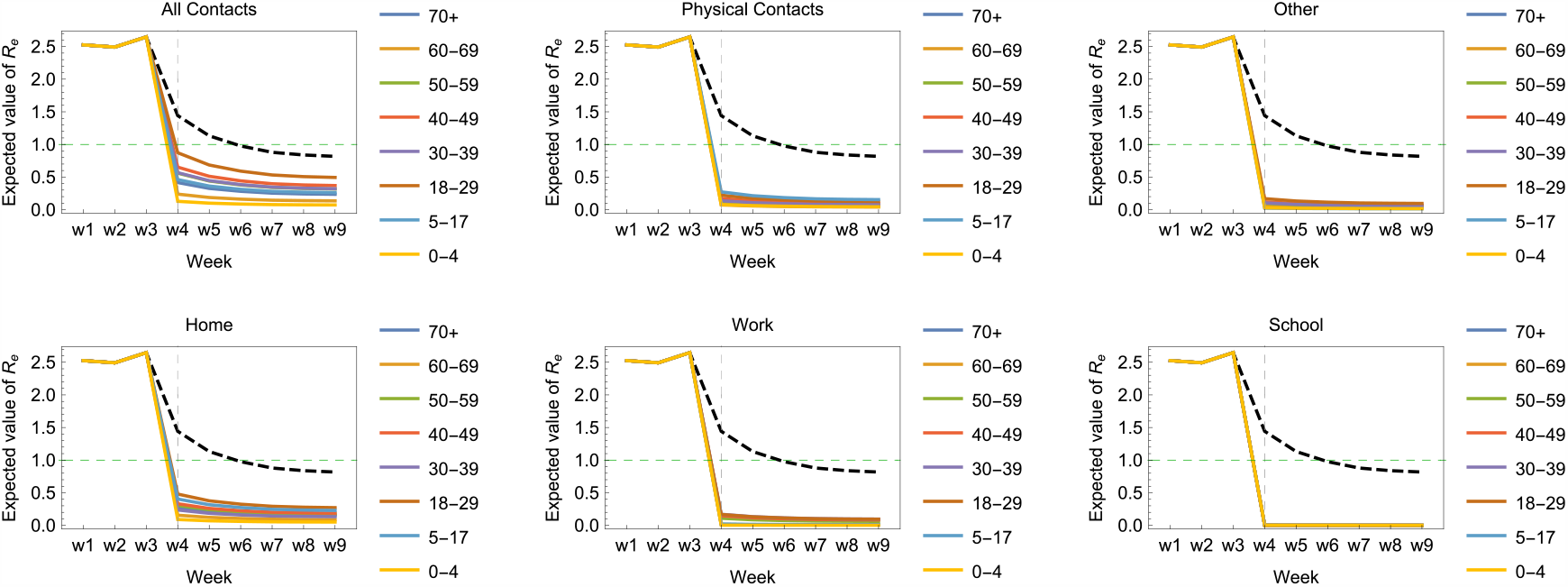
The expected value of the reproduction number as calculated by the Eq. (3.9) for London over nine weeks beginning from the 22nd of February 2020 for different age groups in different social setups. The dashed grey line on each plot shows the 23rd of March 2020 when the lockdown was imposed. Additionally, the black dashed line is the value of 𝔼(ℛ_*e*_) when there is only contact tracing involved and no social mixing (i.e. assuming *C*_*λσ*_ = 1) in Eq. (3.9).

In estimation of 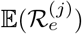 we assumed that the average number of contacts between different age groups in London is roughly the same as the whole UK. We acknowledge that Londoners form only 16.5% of the participants in CoMIX survey yet for the simplicity we assume the contact behaviour of Londoners is the same with the rest of the country. We also assumed perfect tracing in this study, the estimation of *θ* is the subject of upcoming work.

## 4 Conclusions

In this study we proposed an SEIR model with contact tracing and social mixing. We have shown that the effective reproduction number is proportional to dominant eigenvalue of the contact matrix, a result which is also derived in [21] using another method. In Eq.(2.17) we have derived contact tracing via parameters *ϕ* (probability of contacts of a reported being traced) and *α* (fraction of untraced cases who are reported) and showed that the critical value of the required tracing to keep the effective reproduction number below one is proportional to *αϕ*. We also derived the effective reproduction number when there is higher order tracing in Eq. (3.4), in which case there is a linear contribution to the effective reproduction number given by *ϕβ*_*m*_*T*_*m*_ where *β*_*m*_ is the transmissibility of those infected individuals who are monitored and *T*_*m*_ is the time between symptom onset and isolation for the monitored infectious individuals. In the case of the COVID-19 we can assume that *β*_*m*_ ≈ *β*, yet to have an effective contact tracing and lower values of ℛ_*e*_ it is important to keep *T*_*m*_ at its minimum - this of course requires an effective human capital to isolate the monitored cases immediately upon developing symptoms.

We furthermore gave a formula in Eq. (3.9) to estimate the expected value of the effective reproduction number 𝔼(ℛ_*e*_) on a given week. This formula estimates the average reproduction number by using the serial interval of the COVID-19 and the fraction of traced and untraced reported cases. We used this formula to estimate the 𝔼(ℛ_*e*_) in London, UK over nine weeks beginning from the 22nd of February 2020. We used the proportionality of the effective reproduction number to social mixing matrices to calculate the 𝔼(ℛ_*e*_) for six setups and six age groups. While our method in estimating the 𝔼(ℛ_*e*_) is accurate and matches with the values of other studies (such as [25] for London) it is important to improve this calculation by using the formula presented in Eq. (3.4). This equation depends on the infectivity parameters, such as *β, T*_*s*_ and *T*_*a*_, and they can be estimated by methods such as Markov chain Monte Carlo (MCMC) (see for instance [13]). This task is the subject of our upcoming work.

From our model and analysis it is evident that contact tracing and social mixing are both playing important roles in controlling the value of the effective reproduction number (see also [22, 23, 24, 28, 29, 30]). While there is an increasing demand to open up the society since the value of ℛ_*e*_ is below the critical value of one it is important for policymakers to note that the value of ℛ_*e*_ can rebound to higher values should the easing of the restrictions cause higher number of contacts between individuals. In fact, in opening up the society one must suppress the opportunities for transmission. From our model this can be done by firstly, putting an effective contact tracing measure in place, that is to trace, monitor and isolate the contacts of reported cases without time-wasting and secondly, monitoring the number of contacts between different age groups and the fraction of the reported cases classified by their age *α*_*λ*_ to prioritise contacts tracing by age groups when there is a lack of health workers.

## Data Availability

Not Applicable.

## Acknowledgements

The author would like to thank Mikhail Goykhman for useful feedbacks and discussion.

